# Age-Specific SARS-CoV-2 Infection Fatality and Case Identification Fraction in Ontario, Canada

**DOI:** 10.1101/2020.11.09.20223396

**Authors:** David N. Fisman, Steven J. Drews, Ashleigh R. Tuite, Sheila F. O’Brien

**Affiliations:** Dalla Lana School of Public Health, University of Toronto, Toronto, Ontario, Canada; Canadian Blood Services, Ottawa, Alberta, Canada; Canadian Blood Services Ontario and Edmonton, Alberta, Canada

**Author notes:** **Address reprint requests and correspondence to:** David Fisman, MD MPH FRCP(C), Room 686, 155 College Street, Toronto, Ontario, M5T 3M7.

## Abstract

**Background:** SARS-CoV-2 is a novel pandemic pathogen that displays great variability in virulence across cases. Due to limitations in diagnostic testing only a subset of infections are identified. Underestimation of true infections makes calculation of infection fatality ratios (IFR) challenging.

Seroepidemiology allows estimation of true cumulative incidence of infection in populations, for estimation of IFR.

**Methods:** Seroprevalence estimates were derived using retention samples stored by Canadian Blood Services in May 2020. These were compared to non-long-term care-linked case and fatality data from the same period. Estimates were combined to generate IFR and case identification fraction estimates.

**Results:** Overall IFR was estimated to be 0.80% (0.75 to 0.85%), consistent with estimates from other jurisdictions. IFR increased exponentially with age from 0.01% (0.002 to 0.04%) in those aged 20-29 years, to 12.71% (4.43 to 36.50%) in those aged 70 and over. We estimated that 5.88 infections (3.70 to 9.21) occurred for every case identified, with a higher fraction of cases identified in those aged 70 and older (42.0%) than those aged 20-29 (9.4%). IFR estimates in those aged 60 and older were identical to pooled estimates from other countries.

**Conclusions:** To our knowledge these are the first Canadian estimates SARS-CoV-2 IFR and case identification fraction. Notwithstanding biases associated with donor sera they are similar to estimates from other countries, and approximately 80-fold higher than estimates for influenza A (H1N1) during the 2009 epidemic. Ontario’s first COVID-19 pandemic wave is likely to have been accurately characterized due to a high case identification fraction.

## Background

The COVID-19 pandemic has caused over one million counted deaths, and over 40 million counted cases, as of late October 2020 (1). However, a notable feature of the pandemic has been the degree to which cases are under-identified, particularly in younger age groups that have less severe disease (2). Sero-epidemiological studies provide a means of estimating the true burden of infection in populations, and allow calculation of the infection-fatality ratio (the ratio of deaths to infections), an index of virulence that is less likely to fluctuate based on testing capacity than is the case fatality ratio (3).

While purposive, population-based serosurveys are ideal (3-6), they require substantial resources and organization, and may be particularly challenging in the context of restricted movement and contact during a pandemic. Seroprevalence estimates derived from blood donor populations provide an efficient, rapid means for estimating population seroprevalence of infection among adults (3, 7). While blood donors may differ in some respects from the general population (in that they are pre-screened for febrile illness, and less likely to have chronic medical conditions) (8), they may nonetheless provide rapid information about true burden of infection in multiple geographic regions, age groups, and time periods. In addition to providing information on the true underlying cumulative incidence of infection in populations, such data can also be used to estimate the degree to which cases are under-identified in populations (mathematically equivalent to test probability per case) (9).

We combined seroprevalence data from blood with individual-level data case data generate estimates of infection fatality, fraction of cases identified and propensity of cases to undergo testing in Canada’s largest province.

## Methods

### Data sources

Ontario is Canada’s most populous province, with a current population of 14.7 million (10). The province imported cases of SARS-CoV-2 infection from Iran and China in January and February, but available phylogenetic data suggest that eastern Canada’s epidemic was seeded by cases from New York State and Europe in February 2020 (11-13). Ontario has 34 local public health units; each is responsible for local case investigation and uploading of case information the provincial surveillance and case management system (the Integrated Public Health Information System or “iPHIS” (14). Ontario’s case definition for a confirmed case of SARS-CoV-2 requires a positive laboratory test using a validated nucleic acid amplification test, including real-time PCR and nucleic acid sequencing (15). Information on underlying health conditions, long term care residence, age (by 10-year age category), and outcome (fatal or nonfatal) are captured in the iPHIS system.

Blood donation and transfusion in Canada, outside Quebec, are provided by Canadian Blood Services, which initiated serological testing on “retention” samples left over from routine testing between May 9 and June 8, 2020. Blood donation is carried out in large cities and smaller urban areas in Canada and blood donors represent a healthy subset of the Canadian population due to exclusion criteria for donation. Donors who have a history of COVID-19 must wait at least 2 weeks after their symptoms have resolved before they can donate. Retention samples are tubes of blood collected in case supplementary testing is needed after initial infectious disease screening is performed; only around 20% of retention samples are used; the 80% of retention samples not needed for operational testing were aliquoted and frozen at −20°C or colder. All plasma samples were tested using the Abbott Architect SARS-CoV-2 IgG assay (chemiluminescent microparticle immunoassay (CMIA)). Testing was conducted at Canadian Blood Services in Ottawa.

### Estimation of Seroprevalence

To make inferences about the general population, weighting factors were applied based on the donor’s residential Forward Sortation Area, age group and sex. Data were weighted based on Statistic Canada estimates (16). For FSAs with few donors, several FSAs were combined, generally to include at least 500 donors. For data with no FSA recorded weighting was based on FSA of the blood centre. Seroprevalence was calculated as the number of positive samples divided by all samples tested.

### Calculation of Infection Fatality Ratios and Case Identification Fractions

Both seroprevalence data and COVID-19 related deaths likely reflect infections that occurred prior to the date of measurement; the Infectious Disease Society of America regards 14 days as representing a sufficient delay for seropositivity to be identified after symptom onset (17). Our seropositivity estimates were assumed to represent seropositivity at the midpoint of the test period (May 24, 2020); as such comparator cases were those that occurred up to May 9, 2020. We used cases and deaths occurring in individuals with symptom onset up to May 9, 2020 for calculation of infection fatality ratios and case identification fractions, regardless of when deaths occurred. The mean delay between case onset and reporting was 6 days; consequently, we used test volumes to May 15, 2020 for estimation of propensity for testing among cases. Infection fatality ratios were estimated as age-specific deaths divided by total age-specific infections, calculated as age-specific population in Ontario multiplied by age-specific seroprevalence. Case identification fractions were estimated as cases divided by total age-specific infections. Population denominators were derived from Statistics Canada census data (10).

A large fraction of identified infections and deaths during Ontario’s first wave occurred in the long-term care system; these outbreaks were highly clustered, likely had a distinct epidemiology from transmission in the community and were also associated with an extremely high case fatality ratio (18). Furthermore, individuals in long term care facilities would not be included in the eligible blood donor population. As such, cases and deaths in long-term care residents were excluded from analysis.

### Confidence Intervals

Confidence intervals for seroprevalence estimates were calculated assuming a binomial distribution. For infection fatality ratios and case identification fractions, ratios were converted to their logits, and the standard error of logits was estimated as

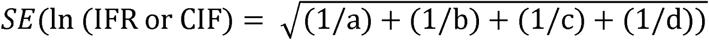

### Sensitivity Analyses and Global Comparison

We performed two sensitivity analyses to account for possible differences in disease risk and risk of severe outcomes among blood donors. In the first, analyses were repeated based on changing the population denominator to reflect the fact that only a subset of Canadians are eligible to donate blood. Cases with record of a chronic medical condition that might be considered a contraindication to blood donation (including chronic liver disease, chronic renal disease, tuberculosis, cardiovascular disease, neurological disease, malignancy, diabetes, hematological disease), travel identified as a risk factor for COVID-19 infection, active injection drug use, incarceration in a correctional facility, or pregnancy were excluded. To make fair comparisons based on a population denominator that reflected these exclusions, we considered the blood donor population to be the 51.9% of the population aged 17 and older who would be eligible to donate blood (8).

As Meyerowitz-Katz has noted (3), in jurisdictions that have performed both purposive serosurveys and blood donation-based serosurveys in parallel (for example in England and Denmark) (19-23) seroprevalence has been approximately 2-fold higher in blood donor surveys, possibly reflecting increased COVID-19 risk tolerance in individuals who are prepared to travel and donate blood during a pandemic (3). To account for this possibility, we repeated our IFR estimates using seroprevalence estimates reduced by 50% in a second sensitivity analysis.

### Global Comparison

We extracted observed and modeled (via meta-regression techniques) estimates for log-transformed IFR by age from the recent meta-analysis of Levin et al. using WebPlotDigitizer (24). We plotted Ontario IFR estimates derived above against estimates from Belgium, Sweden, Spain and Geneva, Switzerland, and also against the fitted meta-regression curve from Levin (3). We assumed a mean age of 75 in the 70 and over group in Ontario, and also assumed that the mean age in other groups was at the mid-point of the age-group (e.g., the 20 to 29 year age group was assumed to have a mean age of 25). All analyses were performed using Stata SE version 15.0 (College Station, Texas).

### Ethics Statement

The study received ethics approval from the Research Ethics Boards at the University of Toronto and Canadian Blood Services. None of the authors has a conflict of interest associated with this work.

## Results

Serological testing was performed on retention samples from 19,387 Ontario blood donors aged 20 and over. Weighted seroprevalence, population sizes, and COVID-19 cases and deaths for the relevant comparator period are presented in **Table 1**. Overall, 0.9% (0.57 to 1.40%) of adults in Ontario were estimated to have been infected with SARS-CoV-2 by May 10, 2020. While seroprevalence was highest in individuals aged 20-29, reported case incidence was highest in individuals aged 50-59; cumulative mortality increased with age from fewer than 1 death per 100,000 among those aged 20-29 to 56 per 100,000 in those aged 70 and older. Case fatality and estimated infection fatality ratios both increased markedly with age; the ratio of CFR to IFR was highest in youngest individuals (11), ranged from 4-6 in middle ages, and was lowest (2.4) in those aged 70 and over.

**Table 1.**
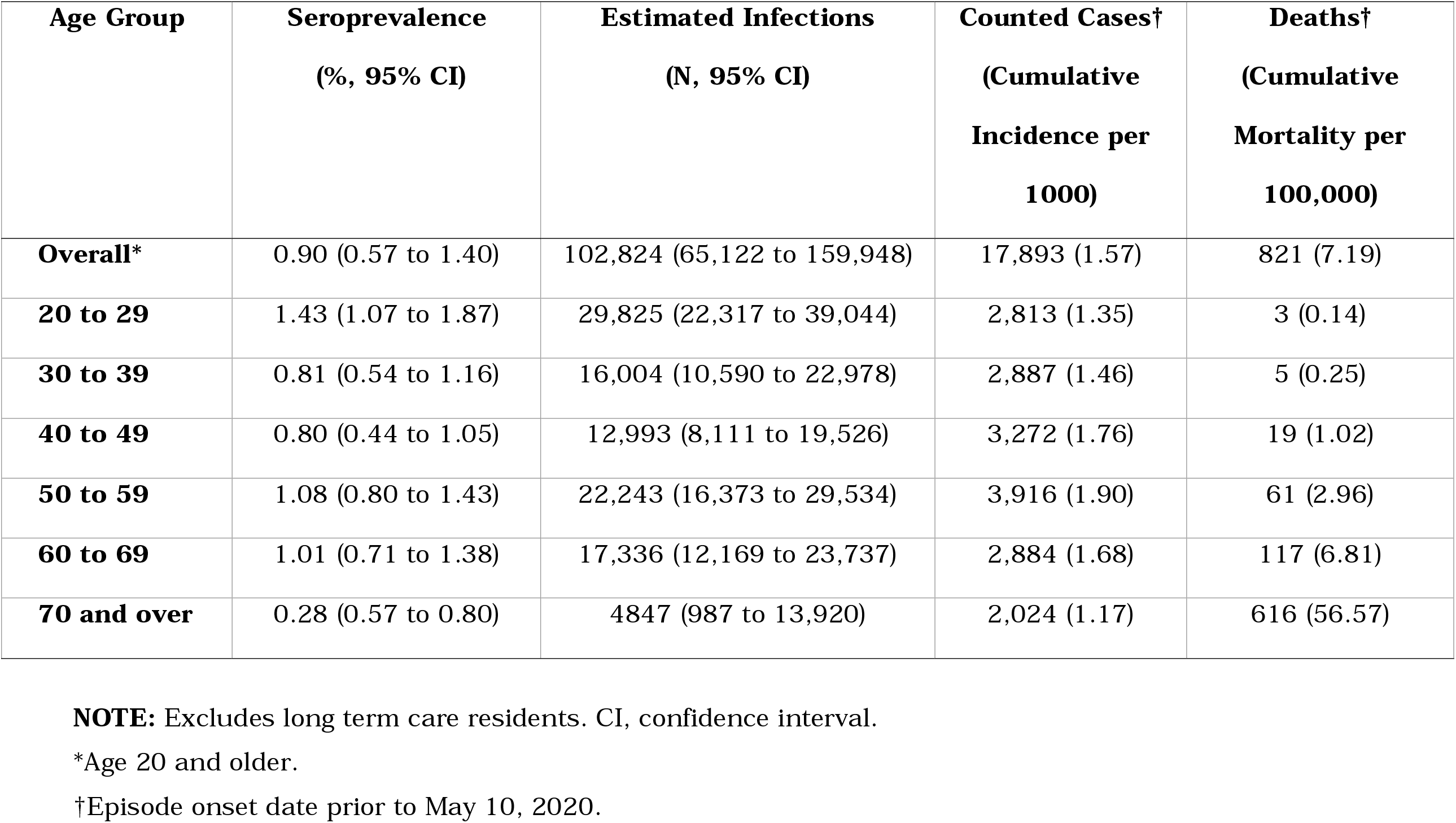
Seroprevalence, Estimated Infections, and Counted COVID-19 Cases and Deaths in Ontario, Canada Prior to May 10, 2020.

Case identification fractions followed a similar pattern, ranging from 9% in youngest individuals to 42% in oldest individuals (**Table 2**). Restriction of case data to individuals without a contradiction to blood donation, and commensurate reduction in the size of population denominators, reduced overall infection fatality to 0.25% (95% CI 0.21 to 0.30). By contrast, when we assumed that seroprevalence was overestimated among blood donors (25), overall infection fatality ratio rose to 1.62% (95% CI 1.51 to 1.73%).

**Table 2.**
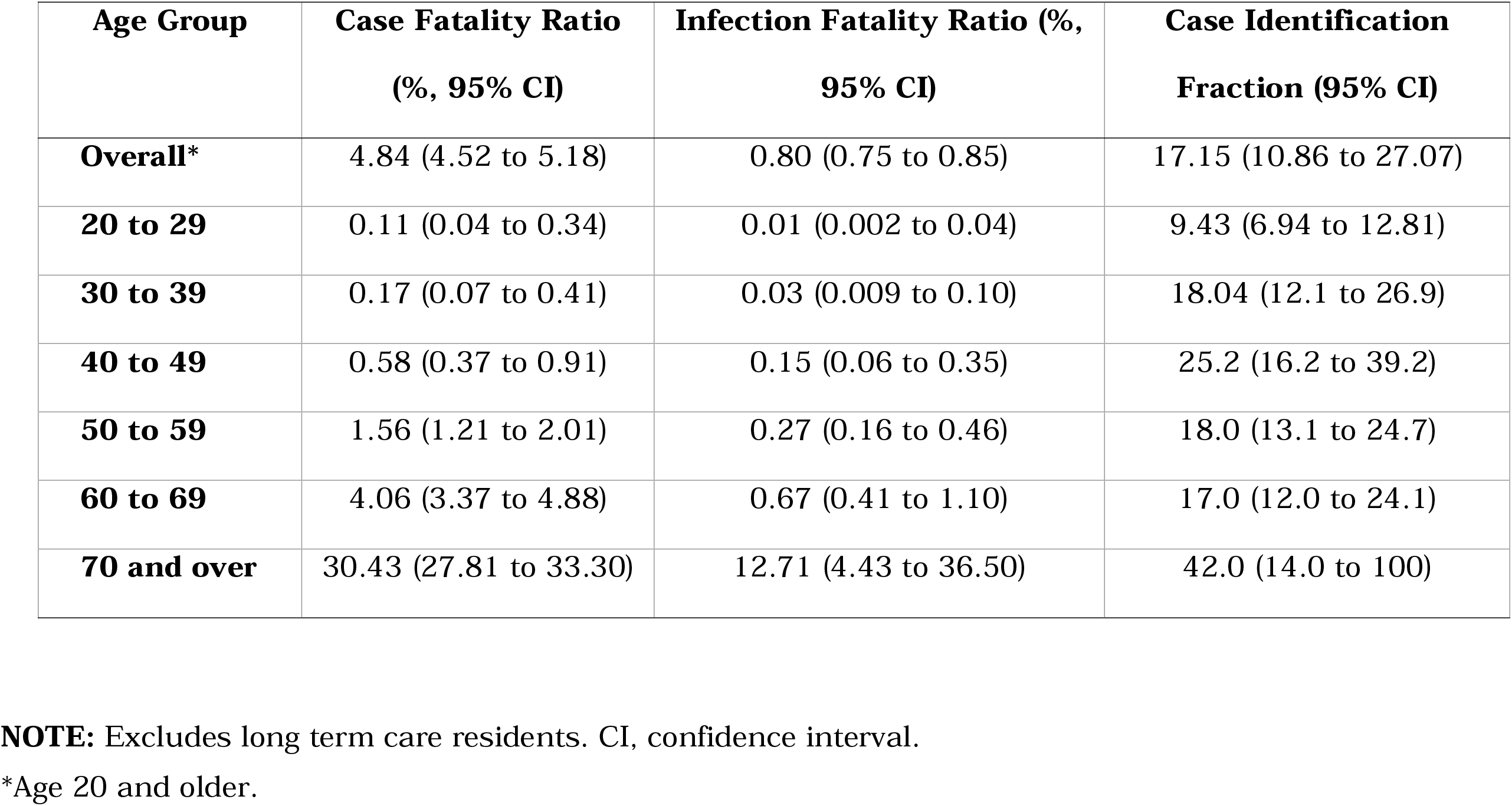
Case Fatality, Infection Fatality and Case Identification Fraction for SARS-CoV-2 Infection in Ontario, Canada Prior to May 10, 2020.

We placed Ontario’s infection fatality ratio estimates in context by plotting them against estimates from four European countries, and fitted IFR curves derived from Levin (3). For older age groups, Ontario estimates fit well with those from Europe. However, IFR estimates for younger individuals were higher than those estimated for comparator countries (**Figure 1**).

**Figure 1.**
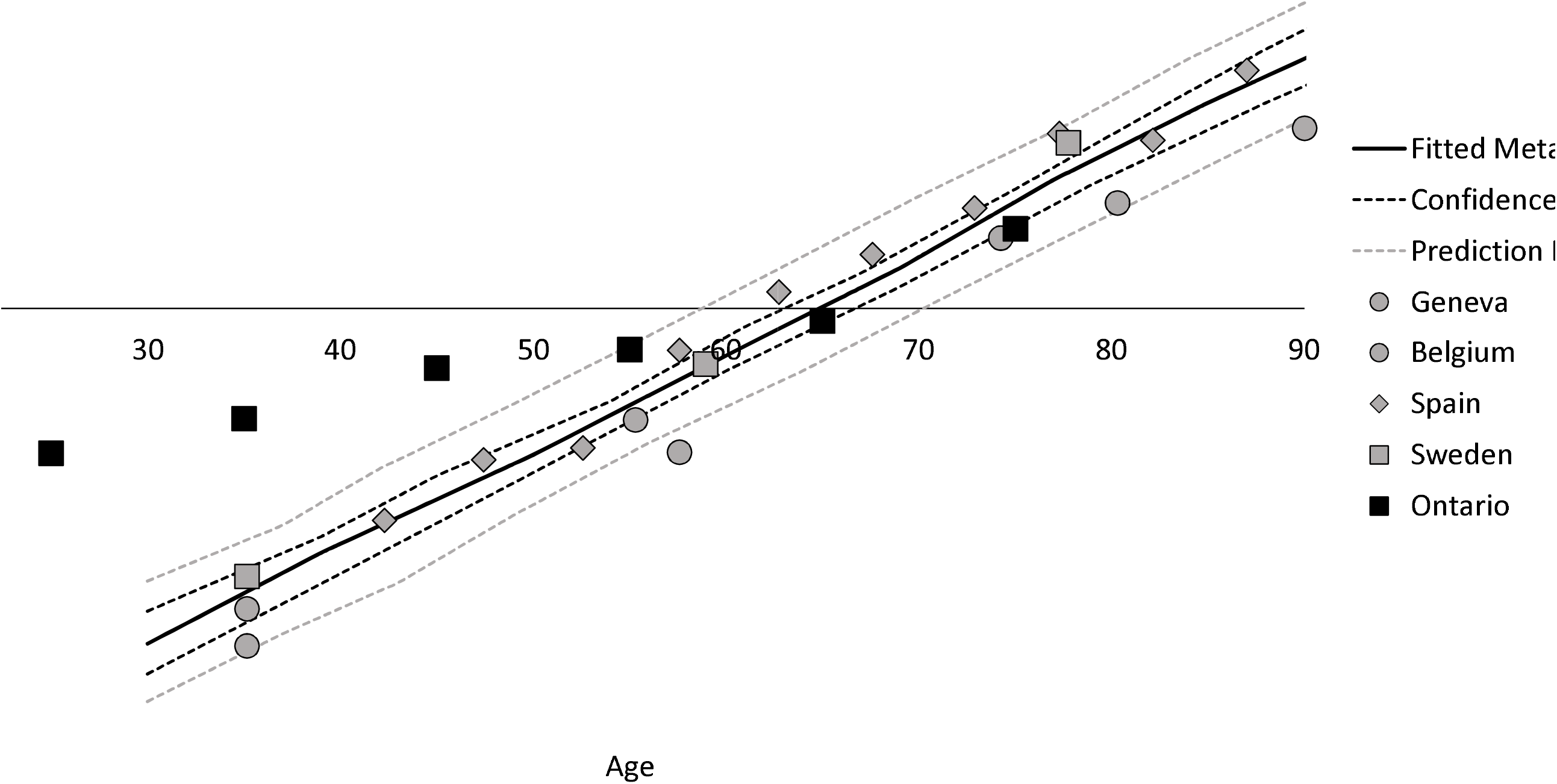
Comparison of Age-Specific Infection Fatality Ratio Estimates in Ontario to Comparator Regions. We overlayed percentage IFR estimates for Ontario (black squares) on a meta-regression figure extracted from Levin et al (3). IFR is plotted on a log_10_ scale (Y-axis); age groups are on the X-axis. It can be seen that Ontario IFR is lower than international estimates in younger individuals, but identical to pooled estimates in those aged 60 and older. Best fit pooled estimate from Levin et al., solid black line; 95% confidence interval, dashed black line; Bayesian prediction interval, dashed gray line.

## Discussion

We evaluated infection fatality ratio due to SARS-CoV-2 infection in Ontario, Canada, using seroprevalence data derived from blood donors. While blood donor-derived seroprevalence estimates are known to have limitations (3, 26, 27), we find the overall IFR estimate in this population to be similar to estimates from a number of other countries. The relatively high quality of Ontario’s case data resources allowed us to avoid overestimation of IFR via exclusion of individuals in long-term care settings, who are likely to experience extreme risk of mortality conditional on COVID-19, and who have experienced outbreak epidemiology in Ontario that is distinct from that seen in the community-dwelling populations from which blood donors are drawn (18, 28).

Even after sensitivity analyses that resulted in exclusion of individuals from the numerator who would have been unable to donate blood due to medical or age contraindications, our resultant lower bound IFR estimate remained similar in order of magnitude to those obtained elsewhere, and also approximately two orders of magnitude higher than IFR estimates published for influenza (29). While adjusting seroprevalence to account for the possibility that it is higher in blood donors than the general population resulted in a predictable increase in IFR, even these upper bound estimates were of similar order of magnitude to estimates published previously (3, 4).

Not unexpectedly, we saw marked increases in estimated IFR by age. Perhaps more surprisingly, although our IFR estimates in older adults were similar to those seen elsewhere, we found that our IFR estimates in younger adults, while very low, were far higher than those from Sweden, Switzerland, Spain and Belgium. The geographically patchy nature of SARS-CoV-2 infection in Ontario may explain this discrepancy. Inasmuch as blood donors are typically more affluent and urban than the population as a whole, and Ontario’s SARS-CoV-2 epidemic has been notably concentrated in lower income areas of the province (30), it may be that our seroprevalence estimates underestimate true seroprevalence in disadvantaged parts of the province. Elevated IFR in older adults also reflects lower seroprevalence estimates in this group, which in turn may reflect self-protective behaviors (e.g., greater compliance with public health measures like social distancing) due to increased perceived risk in this age group (2, 31).

In addition to estimation of IFR, we used seroprevalence data to estimate the fraction of cases identified during the first SARS-CoV-2 wave in Ontario; not unexpectedly, we estimated that a higher fraction of cases was identified among older individuals at greater risk of more severe illness. Our case identification fraction suggests a ratio of unidentified to identified SARS-CoV-2 infections of around 6:1, compatible with data from Connecticut, which in turn appeared to identify cases more efficiently than most other comparator jurisdictions in the United States (9). This high level of case identification could represent efficient targeting of testing given the equivalent SARS-CoV-2 testing per capita in Canada and the United States in May 2020 (32). However, differences between donor and non-donor populations may account for these differences (for example, if blood donors are more drawn systematically from lower risk populations).

Like any observational study, ours is subject to limitations. Issues of generalizability related to the use of blood donor samples, rather than a purposive, random serosurvey are noted above. Seroreversion is increasingly recognized as an issue that artificially decreases seroprevalence for SARS-CoV-2 (33), but the fact that our serosurvey was completed shortly after the first wave of infection in Ontario likely makes this less of an issue here. Lastly, our dataset does not include children, whose role in the dynamics of the SARS-CoV-2 pandemic remains controversial. Nonetheless, we are able to conclude that the infection fatality ratio from SARS-CoV-2 infection in Ontario is similar to IFR reported in other countries, age-specific IFR in older adults is similar to published estimates from elsewhere, and that case identification fraction is likely linked to disease severity, and is higher in Ontario than in other North American jurisdictions where such data have been reported.

## Data Availability

Please contact Dr. Fisman for information on data availability.

## Notes

The research was supported by a grant to DNF from the Canadians Institutes for Health Research (2019 COVID-19 rapid researching funding OV4-170360).

### Competing Interest Statement

The authors have declared no competing interest.

### Funding Statement

Funded by the Canadian Institutes for Health Research.

### Author Declarations

Approved by the Research Ethics Boards of the University of Toronto and Canadian Blood Services

